# Modeling the combined effect of digital exposure notification and non-pharmaceutical interventions on the COVID-19 epidemic in Washington state

**DOI:** 10.1101/2020.08.29.20184135

**Authors:** Matthew Abueg, Robert Hinch, Neo Wu, Luyang Liu, William Probert, Austin Wu, Paul Eastham, Yusef Shafi, Matt Rosencrantz, Michael Dikovsky, Zhao Cheng, Anel Nurtay, Lucie Abeler-Dörner, David Bonsall, Michael V. McConnell, Shawn O’Banion, Christophe Fraser

## Abstract

Contact tracing is increasingly being used to combat COVID-19, and digital implementations are now being deployed, many of them based on Apple and Google’s Exposure Notification System. These systems are new and are based on smartphone technology that has not traditionally been used for this purpose, presenting challenges in understanding possible outcomes. In this work, we use individual-based computational models to explore how digital exposure notifications can be used in conjunction with non-pharmaceutical interventions, such as traditional contact tracing and social distancing, to influence COVID-19 disease spread in a population. Specifically, we use a representative model of the household and occupational structure of three counties in the state of Washington together with a proposed digital exposure notifications deployment to quantify impacts under a range of scenarios of adoption, compliance, and mobility. In a model in which 15% of the population participated, we found that digital exposure notification systems could reduce infections and deaths by approximately 8% and 6%, effectively complementing traditional contact tracing. We believe this can serve as guidance to health authorities in Washington state and beyond on how exposure notification systems can complement traditional public health interventions to suppress the spread of COVID-19.

## Introduction

The COVID-19 pandemic has brought about tremendous societal and economic consequences across the globe, and many areas remain deeply affected. Due to the urgency and severity of the crisis, the poorly understood long-term consequences of the virus, and the lack of certainty about which control measures will be effective, many approaches to stopping or slowing the virus are being explored.

In seeking solutions to this problem, many technology-based non-pharmaceutical interventions have been considered and deployed (*1*), including data aggregation to track the spread of the disease, GPS-enabled quarantine enforcement, AI-based clinical management, and many others.

Contact tracing, driven by interviews of infected persons to reveal their interactions with others, has been a staple of epidemiology and public health for the past two centuries (*2*). These human-driven methods have been brought to bear against COVID-19 since its emergence, with some success (*3*). Unfortunately, owing in part to the rapid and often asymptomatic spread of the virus, these efforts have not been successful in preventing a global pandemic. Further, as infections have reached into the millions, traditional contact tracing resources have been overwhelmed in many areas (*4*) (*5*). Given these major challenges for traditional contact tracing, technology-based improvements are being explored, with particular focus on the use of smartphones to detect exposures to others carrying the virus.

Smartphone apps may approximate pathogen exposure risk through the use of geolocation technologies such as GPS, and/or via proximity-based approaches using localized Radio Frequency (RF) transmissions like Bluetooth. Location-based approaches attempt to compare the places a user has been with a database of high-risk locations or overlaps with infected people (*6*), while proximity-based approaches directly detect nearby smartphones that can later be checked for “too close for too long” exposure to infected people (*7*). In either approach, users who are deemed to be at risk are then notified, and in some implementations, health authorities also receive this information for follow-up.

Due to accuracy and privacy concerns, the majority of contact tracing proposals have avoided the location signal and focused on a proximity-based approach, such as PEPP-PT (*8*) and NSHX (*9*). Further privacy safeguards may be achieved by decentralizing and anonymizing important elements of the system, as in DP-3T(*10*) and Apple and Google’s Exposure Notifications System (ENS) (*11*). In these approaches, the recognition of each user’s risk level can take place only on the user’s smartphone, and server-side knowledge is limited to anonymous, randomized IDs.

Technological solutions in this space have never been deployed at scale before, and their effectiveness is unknown. There is an acute need to understand their potential impact, to establish and optimize their behavior as they are deployed, and to harmonize them with traditional contact tracing efforts. Specifically we will examine these issues in the context of ENS, which is currently being adopted by many countries (*12*).

There are many variables to consider when characterizing the behavior of any system of this type. Technology-dependent parameters, such as those needed to convert Bluetooth signal strength readings to proximity (*13*) (*14*), vary from device to device and require labor-intensive calibration. They will not be discussed in this paper. Here we seek to explore the general conditions and public health backdrop in which an ENS deployment may exist, and the policy characteristics that can accompany it.

In order to improve our understanding of this new approach, we employ individual-based computational models, also known as agent-based models, which allow the exploration of disease dynamics in the presence of complex human interactions, social networks, and interventions (*15*). This technique has been used to successfully model the spread of Ebola in Africa (*16*), malaria in Kenya (*17*), and influenza-like illness in several regions (*18*) (*19*), among many others. In the case of COVID-19, the OpenABM-Covid19 model by Hinch et a.l (*20*) has already been used to explore smartphone-based interventions in the United Kingdom. This model seeks to simulate individuals and their interactions in home, work, and community contexts, using epidemiological and demographic parameters as a guide.

In this work, we adapt the OpenABM-Covid19 model to simulate the ENS approach and apply it to data from Washington state in the United States in order to explore possible outcomes. We use data at the county level to match the population, demographic, and occupational structure of the region, and calibrate the model with epidemiological data from Washington state and Google’s Community Mobility Reports for a time-varying infection rate (*21*). Similar to Hinch et al., we find that digital exposure notification can effectively reduce infections, hospitalizations, and deaths from COVID-19 at all levels of participation. We extend the findings by Hinch et al. to show how digital exposure notification can be deployed concurrently with traditional contact tracing and social distancing to suppress the current epidemic and aid in various “reopening” scenarios. We believe the demographic and occupational realism of the model and its results have important implications for the public health of Washington state and other health authorities around the world working to combat COVID-19.

## Methods

### Modeling individual interactions and COVID-19 epidemiology

To model the combined effect of digital exposure notification and other non-pharmaceutical interventions (NPIs) in Washington state, we use a model first proposed by Hinch et al. (*20*), who have also made their code available as open source on GitHub (*22*). OpenABM-Covid19 is an individual-based model that models interactions of synthetic individuals in different types of networks based on the expected type of interaction (Fig. 1). Workplaces, schools, and social environments are modeled as Watts--Strogatz small-world networks (*23*), households are modeled as separate fully connected networks, and random interactions, such as those on public transportation, are modeled in a random network. The networks are parameterized such that the average number of interactions matches the age-stratified data in (*24*). Contacts between synthetic individuals in those interaction networks have the potential for transmission of the virus that causes COVID-19 and are later recalled for contact tracing and possible quarantine.

**Fig. 1.**
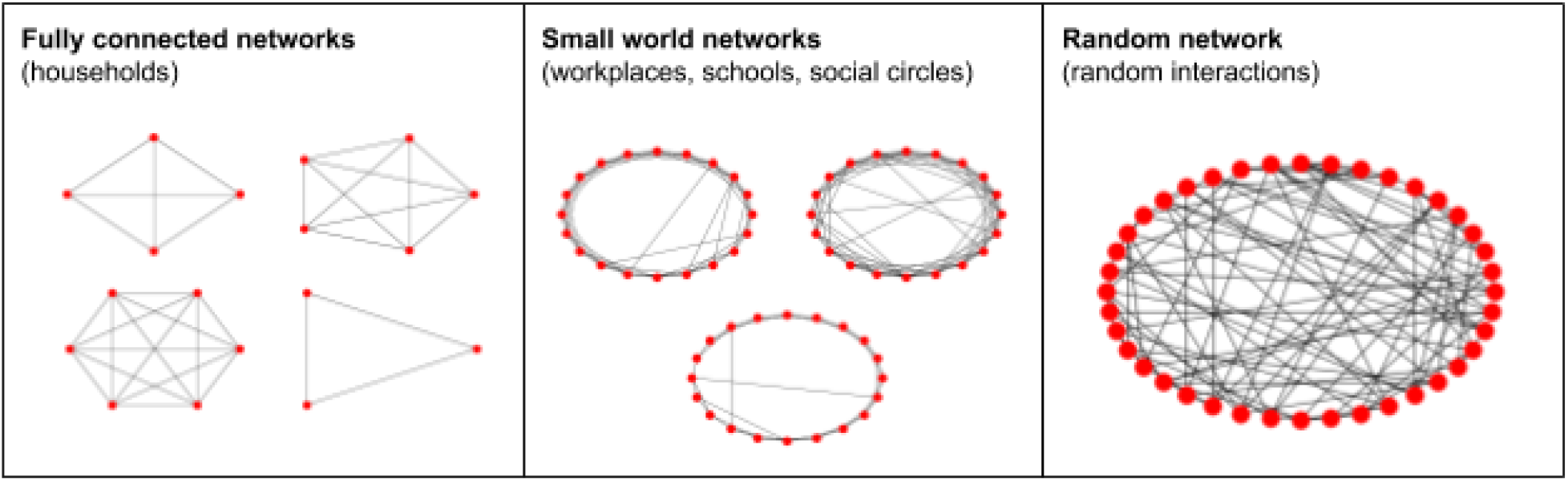
Examples of fully connected, Watts–Strogatz small-world, and random networks that define interactions among synthetic agents in households, workplaces, schools, social circles, and random settings.

While the original model by Hinch et al. (*22*) included a single occupation network for working adults, we extend this to support multiple networks for workplace heterogeneity. This is motivated by increasing evidence that workplace characteristics play an important role in the spread of SARS-CoV-2, such as having to work in close physical proximity to other coworkers and interacting with the public. Baker et al. found that certain U.S. working sectors experience a high rate of SARS-CoV-2 exposure, including healthcare workers, protective services (e.g., police officers), personal care and services (e.g., child care workers), community and social services (e.g., probation officers) (*25*). As another example, the Centers for Disease Control and Prevention (CDC) has issued specific guidance to meat and poultry processing workers due to the possible increased exposure risk in those environments (*26*). Therefore, we model each individual industry sector as its own small-world network and parameterize it with real-world data such as the sector size and interaction rates.

In OpenABM-Covid19, transmission between infected and susceptible individuals through a contact is determined by several factors, including the duration since infection, susceptibility of the recipient (a function of age), and the type of network where it occurred (home networks assume a higher risk of transmission due to the longer duration and close proximity of the exposure). Individuals progress through stages of susceptible, infected, recovered, or deceased. In this model, the dynamics of progression through these stages are governed by several epidemiological parameters, such as the incubation period, disease severity by age, asymptomatic rate, and hospitalization rate, and are based on the current literature of COVID-19 epidemiology. A complete list of the epidemiological parameters can be found at (*27*) and any modifications to those are described in the subsequent sections and documented in the supplementary materials (Table S1, S2).

### Modeling Washington state

In this work we model the three largest counties in Washington state -- King, Pierce, and Snohomish -- with separate and representative synthetic populations. The demographic and household structure were based on data from the 2010 U.S. Census of Population and Housing (*28*) and the 2012-2016 ACS Public Use Microdata Sample (*29*). We combined Census and Public Use Microdata Sample (PUMS) data using a method inspired by (*30*). For each Census block in Washington state we took distributions over age, sex, and housing type from several marginal tables (called Census Summary tables) and from the PUMS, and combined them into a multiway table using the iterative proportional fitting (IPF) algorithm. We then resampled the households from the PUMS to match the probabilities in the multiway table. The resulting synthetic population in each Census block respects the household structure given by PUMS and matches marginals from the Census Summary tables.

Our synthetic working population was drawn to match the county-level industry sector statistics reported by the U.S. Bureau of Labor Statistics in their Quarterly Census of Employment and Wages for the fourth quarter of 2019 (*31*). We also used a report by the Washington State Department of Health (DOH) containing the employment information of lab-confirmed COVID-19 cases among Washington residents as of May 27, 2020 to parameterize each occupation sector network (*32*). For each sector, we use its lab-confirmed case number weighted by the total employment size as a multiplier factor to adjust the number of work interactions of that occupational network. While the DOH report does not explicitly measure exposure risk for different industries, it is, to the best of our knowledge, the best source of data for confirmed COVID-19 cases and occupations to date. Our model should be refined with better data from future work that studies the causal effect of workplace characteristics on COVID-19 transmission. A complete list of the occupation sectors and interaction multipliers can be found in the supplementary materials (Table S3,S4).

### Modeling interventions

#### Testing and quarantine

In the OpenABM-Covid19 model, if an individual presents with COVID-19 symptoms, they receive a test and are 80% likely to enter a voluntary 7-day isolation with a 2% drop out rate each day for noncompliance. If the individual receives a positive test result, they isolate for a full 14 days from initial exposure with a daily drop out rate of 1 %. Prior to confirmation of the COVID-19 case via a test result, the household members of the voluntarily self-isolating symptomatic individual do not isolate, which is in line with current recommendations by the CDC (*33*). Household quarantines may still occur through digital exposure notification or manual contact tracing, described in the following sections.

#### Digital exposure notification

We simulate digital exposure notification in OpenABM-Covid19 by broadcasting exposure notifications to other users as soon as an app user either tests positive or is clinically diagnosed with COVID-19 during hospitalization. The model recalls the interaction networks of this app user, known as the “index case”, to determine their first-order contacts within the previous 10 days. Those notified contacts are then 90% likely to begin a quarantine until 14 days from initial exposure with a 2% drop out rate each day for noncompliance. See (*22*) for a more comprehensive description of the model.

While the actual ENS allows health authorities to configure notifications as a function of exposure distance and duration, our model does not have the required level of resolution and instead assumes that 80% of all “too close for too long” interactions are captured between users that have the app. (See the supplemental materials for a sensitivity analysis of this parameter.)

The overall effect of digital exposure notification depends on a number of factors that we explore in this work, including the fraction of the population that adopts the app and the delay between infection and exposure notification. As an upper bound on app adoption, we configure the age-stratified smartphone population using data on smartphone ownership from the U.S. from the Pew Research Center (*34*) for ages 20+ and Common Sense Media (*35*) for ages 0-19. Since this data was not available for Washington state specifically we assumed that the U.S. distribution was representative of Washington state residents.

#### Manual contact tracing

We also extend OpenABM-Covid19 to model traditional or “manual” contact tracing as a separate intervention. In contrast to digital exposure notification, human tracers work directly with index cases to recall their contact history without the proximity detection capabilities of a digital app. Those contacts are then given the same quarantine instructions as those traced through the digital app. We configure the simulation such that manual contact tracers have a higher likelihood of tracing contacts in the household and workplace/school networks (100% and 80%, respectively) than for the additional random daily contacts (5%). This is based on the assumption that people will have better memory and ability to identify contacts in the former (e.g., involving family members or coworkers) compared to the latter (e.g., a random contact at a restaurant). Additionally, we configure the capacity of the contact tracing workforce with parameters for workforce size, maximum number of index-case interviews per day, and maximum number of tracing notification calls per day following those interviews. Tracing is initiated on an index case after either a positive test or hospitalization, subject to the capacity in that area. Finally, we add a delay parameter between initiation of manual tracing and finally contacting the traced individuals to account for the processing and interview time of manual tracing.

### Model calibration

Model calibration is the process of adjusting selected model parameters such that the model’s outputs closely match real-world epidemiological data. To calibrate OpenABM-Covid19 for Washington state we use components of a Bayesian SEIR model by Liu et al. (*36*) for modeling COVID-19. They extend the classic SEIR model by allowing the infection rate to vary as a function of human mobility and a latent changepoint to account for unobserved changes in human behavior. We fit that model to Washington state county-level mortality data from *The New York Times* (*37*) and mobility data from the Community Mobility Reports published by Google and publicly available at (*21*). The Community Mobility Reports are created with aggregated, anonymized sets of data from users who have turned on the Location History setting, which is off by default. No personally identifiable information, such as an individual’s location, contacts or movement, is ever made available (*38*). The reports chart movement trends over time by geography, across different categories of places such as retail and recreation, groceries and pharmacies, parks, transit stations, workplaces, and residential. We note that, because of the opt-in nature of this dataset, it may not be representative of the overall population.

We extend the methodology in Liu et al. to model calibration in OpenABM-Covid19 by applying the time-varying infection rate coefficients to the relevant county-specific parameters that guide user interaction levels and disease transmission likelihood. More specifically, the number of daily interactions in the random and occupation networks, *Ri*(*t*) and *Wi*(*t*), are scaled by the mobility coefficient, *m*(*t*) at time step *t*, which is calculated based on the aggregated and anonymized location visits from the Community Mobility Reports. The time-dependent infectious rate, β(t), is scaled by a weighting term, σ(*t*), that depends on how far time step *t* is from a learned changepoint, which is modeled as a negative sigmoid. Both σ(*t*) and *m*(*t*) are learned functions and are described in more detail in (36).

Finally, we use an exhaustive grid search to compute two OpenABM-Covid19 parameters for each county: its initial infectious rate and the infection seed date^1^. The infectious rate is the mean number of individuals infected by each infectious individual with moderate-to-severe symptoms, and can be considered a function of population density and social mixing. The infection seed date is the date at which the county reaches 30 total infections, possibly before the first official cases due to asymptomatic and unreported cases. We pick the parameters where the simulated mortality best matches the actual COVID-19 mortality from epidemiological data, as measured by root-mean-square error (RMSE).

The results of the calibrated models for King, Pierce, and Snohomish counties are shown in Fig. 2. Note that while there is a strong correlation in the predicted and reported incidence, the absolute predicted counts are approximately 6X higher than those that were officially reported. We attribute this difference to the fact that OpenABM-Covid19 is counting all asymptomatic and mild symptomatic cases that may not be recorded in reality. This is approximately consistent with the results of a seroprevalence study by the CDC that estimated that there were 6 to 24 times more infections than official case report data (*39*).

**Fig. 2.**
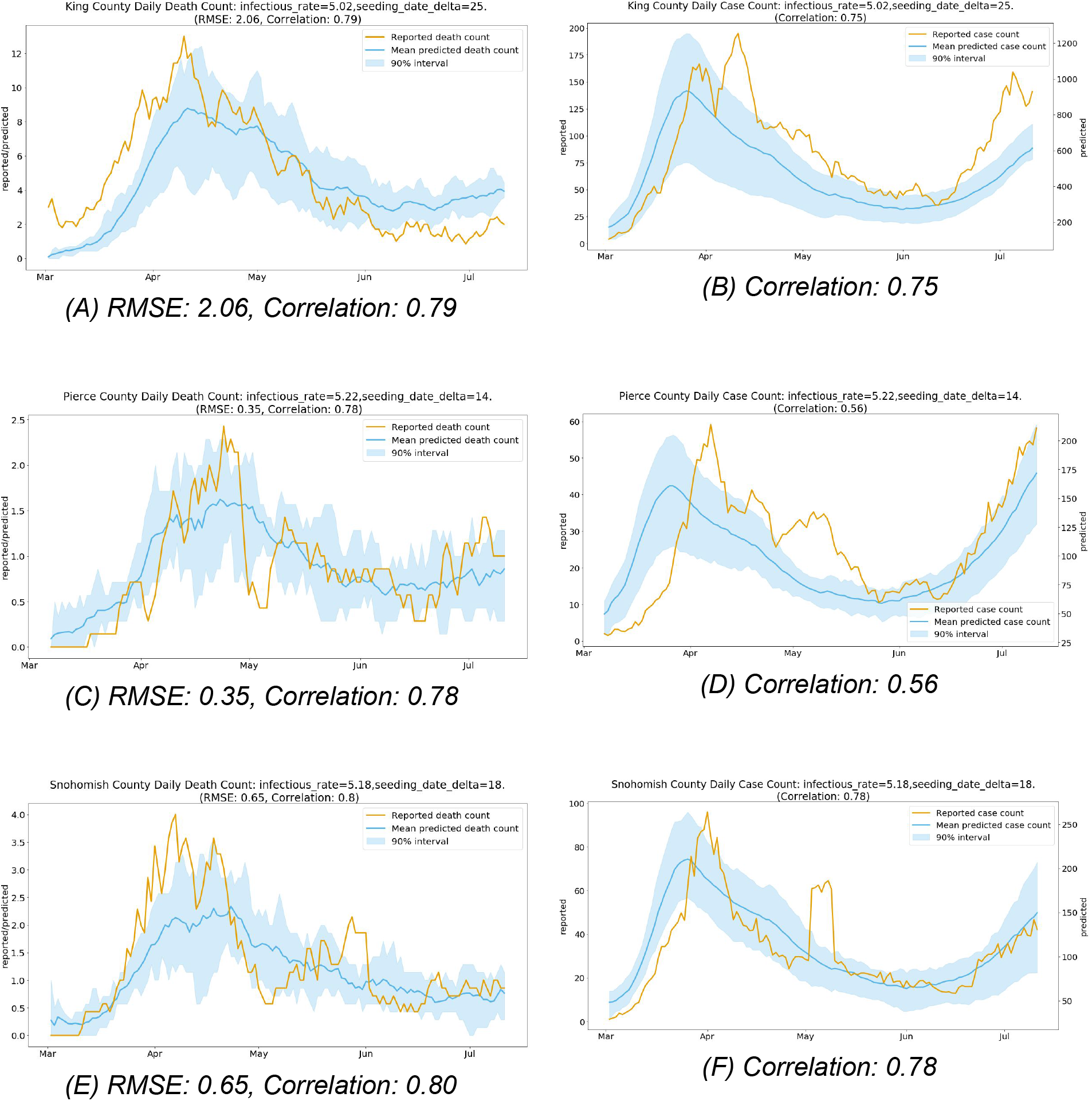
Daily reported and predicted COVID-19 deaths in King County, WA (A), Pierce County, (C), and Snohomish County, WA (E) and daily reported and predicted COVID-19 cases for King County, WA (B), Pierce County, WA (D), and Snohomish County, WA (F).

## Results

In this section we present several forward-looking simulations for Washington state counties by comparing multiple hypothetical scenarios that implement some combination of digital exposure notification, manual contact tracing, or social distancing. Each simulation uses the same calibrated model parameters up to July 11, 2020 at which point the hypothetical interventions are implemented. Beyond this date, each simulation uses the model parameters from the last week of the calibration period, except where explicitly specified as part of the intervention. For each simulated intervention we report the number of infections (daily and cumulative), cumulative number of deaths, number of hospitalizations, number of tests per day, and fraction of the population in quarantine. Each simulation covers 300 consecutive days from March 1, 2020 through Dec 25, 2020, plus the additional calibrated seeding period before March 1. Unless otherwise stated, the reported result is the mean value over 10 runs with different random seeds of infection.

Note that results may be affected by the end date of the simulation because of the time it takes some interventions to have their full effect. We believe that a time horizon of approximately 5 and a half months is long enough to be practically useful for public health agencies who are considering deploying such interventions, but short enough to minimize the long-term uncertainty and effects of externalities such as a vaccine becoming available.

### Digital exposure notification

We first study the effect of a digital exposure notification app at different levels of app adoption -- 15%, 30%, 45%, 60%, and 75% (or all smartphone owners) -- of the population in each county. As a baseline, we compare those results to the “default” scenario without digital exposure notification and assume no change in behavior or interventions beyond July 11, 2020. The results show an overall benefit of digital exposure notification at every level of app adoption (Fig. 3 and 4). When compared to the default scenario of only self isolation due to symptoms, each scenario results in lower overall incidence, mortality, and hospitalizations. Unsurprisingly, the effect on the epidemic is more significant at higher levels of app adoption. An app with 75% adoption reduces the total number of infections by 56-73%, 73-79%, and 67-81% and the number of total deaths by 52-70%, 69-78%, and 63-78% for King, Pierce, and Snohomish counties, respectively. Even at a relatively low level of adoption of 15%, total infections are reduced by 3.9-5.8%, 8.1-9.6%, and 6.3-11.8% and total deaths are reduced by 2.2-6.6%, 11.2-11.3%, and 8.2-15.0% for King, Pierce, and Snohomish counties, respectively.

**Fig. 3.**
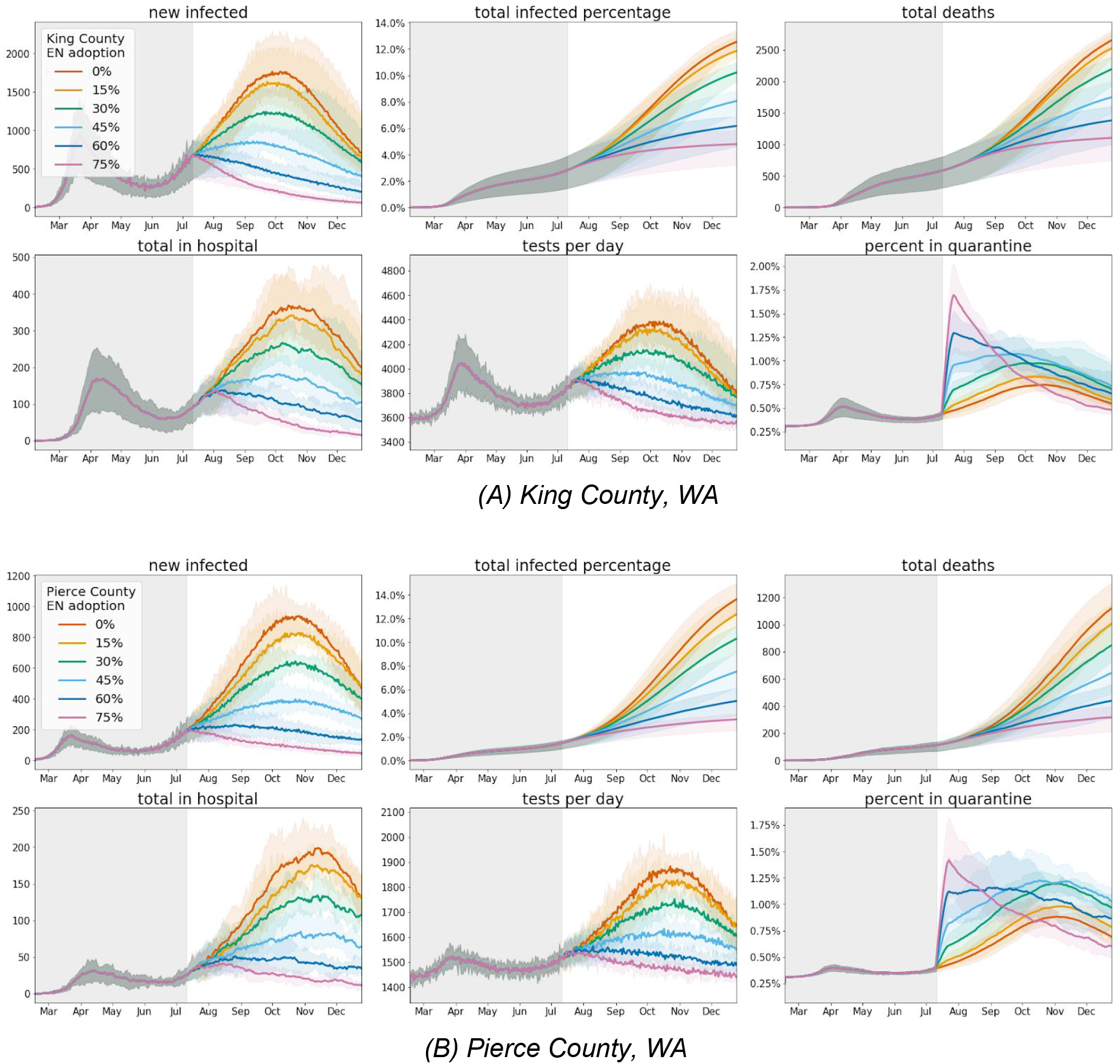
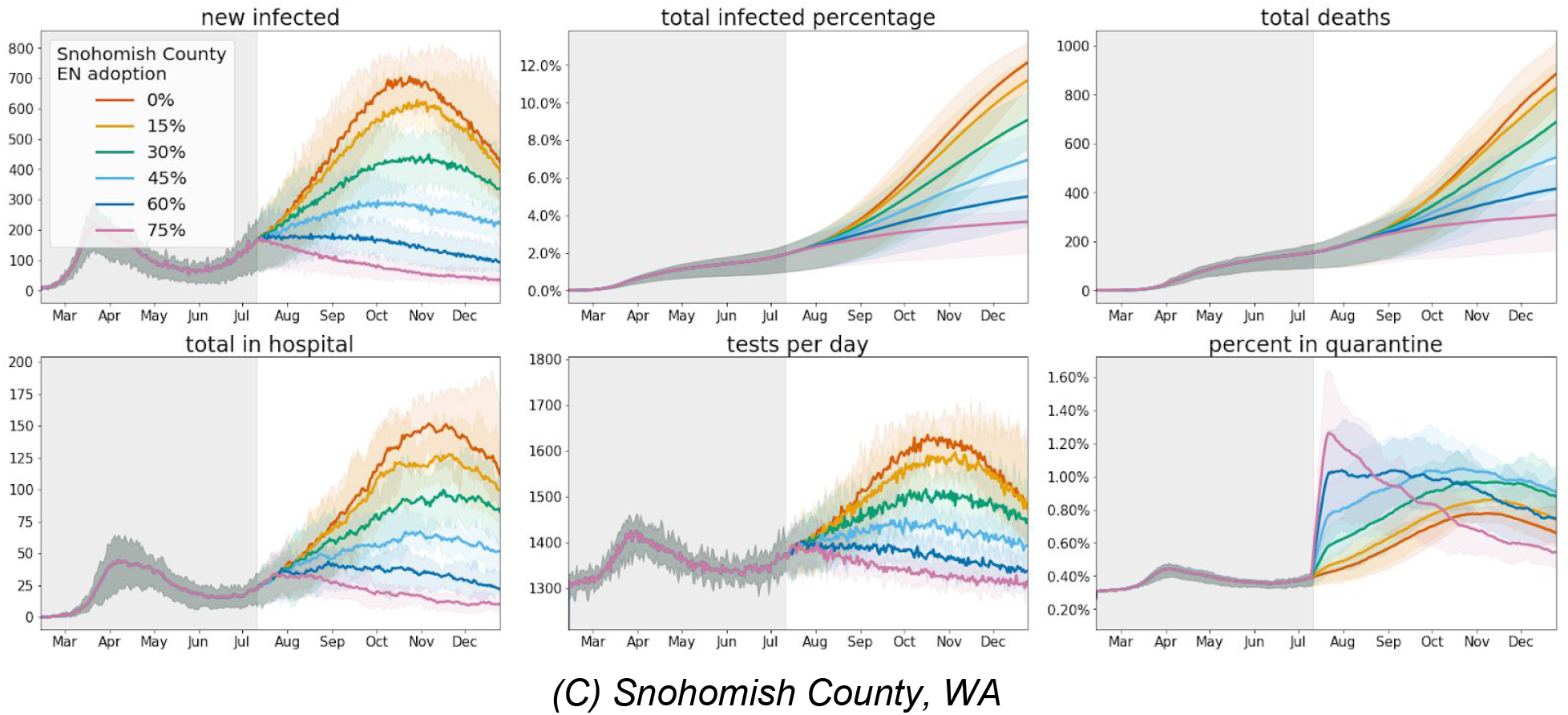
Simulation results for various levels of exposure notification app uptake (among the total population) during 2020, with the app being implemented on July 11, 2020 in (A) King, (B) Pierce, and (C) Snohomish counties. The shaded areas represent the 97.5% confidence intervals.

**Fig. 4.**
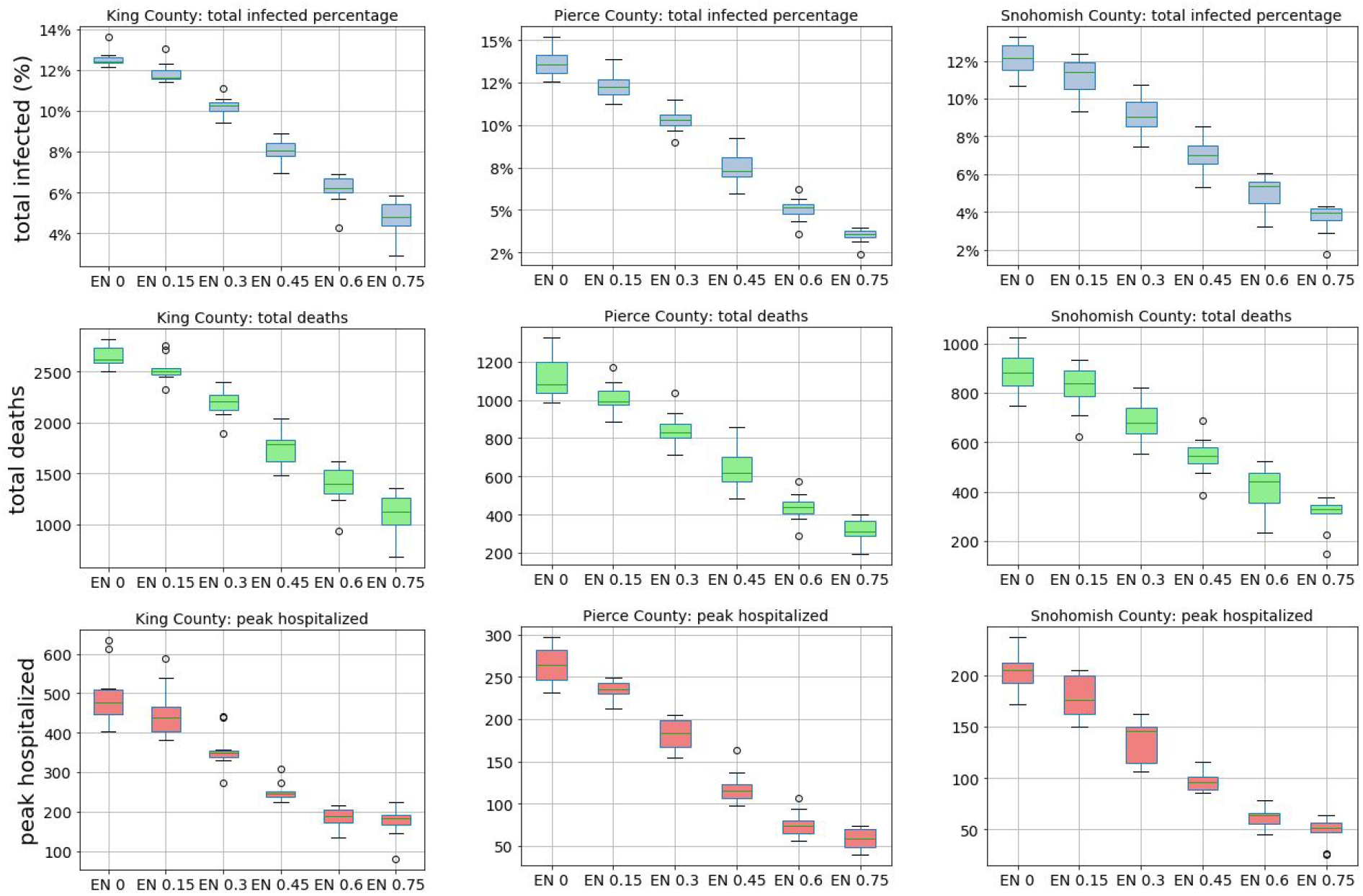
Estimated total infected percentage, total deaths, and peak in hospital (y-axes) of King, Pierce, and Snohomish counties for various levels of exposure notification (EN) app uptake among the population (x-axis) between July 11, 2020 and December 25, 2020. The boxes represent the Q1 to Q3 quartile values with a line at the median. The whiskers show the range of the data (1.5 * (Q3-Q1)) and any outlier points are past the end of the whiskers.

In addition to its effect on the epidemic, we also evaluate the trade-off between exposure notification app adoption and the total number of quarantine events. There is an incentive to minimize the quarantine rate because of the perceived economic and social consequences of stay-at-home orders. At 15% exposure notification app adoption the number of total quarantine events increases by 4.6-6.4%, 6.6-6.8%, and 5.8-10.2% for King, Pierce, and Snohomish counties (Fig. 5). In general, the higher the level of exposure notification adoption the greater the number of total quarantine events, with the exception of very high levels of adoption (60% and 75%) where this number plateaus or even decreases, likely due to the significant effect of the intervention in suppressing the overall epidemic in those scenarios. From another perspective, achieving epidemic control at the price of high initial quarantine is preferable to lower levels of quarantine that are sustained for much longer.

**Fig. 5:**
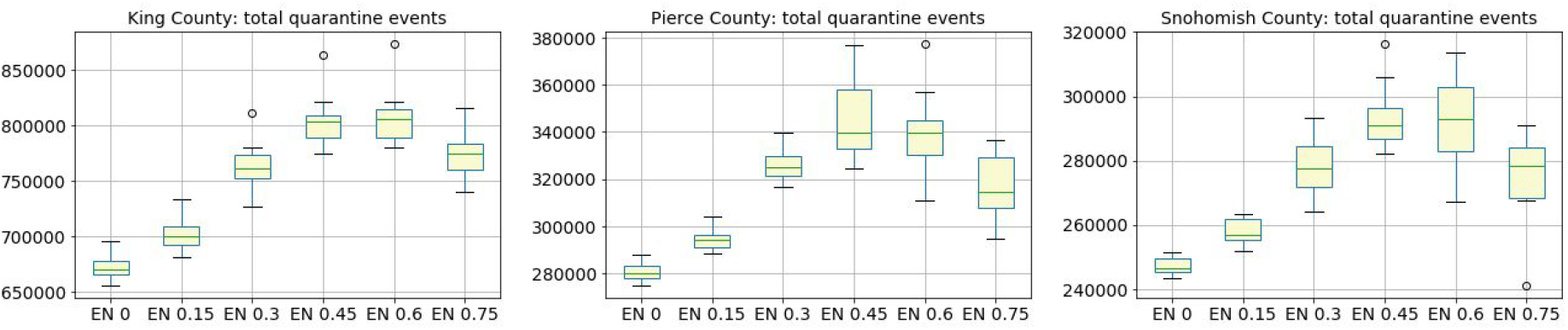
Estimated total quarantine events of King, Pierce, and Snohomish counties for various levels of exposure notification app uptake among the population from July 11, 2020 to December 25, 2020. Note that even for the “default” (0% EN app uptake) scenario there is a non-zero number of quarantine events because this assumes that symptomatic and confirmed COVID-19 positive individuals will self-quarantine at a rate of 80%, even in the absence of an app.

### Manual contact tracing

Next we study the potential impact of manual contact tracing in suppressing the epidemic as a function of the contact tracing workforce size. We refer to the Office of the Governor of WA State that recommends, at minimum, 15 tracers per 100,000 people. Furthermore we use the current staffing rates for King County including all available staffers (105 full-time workers for 2.253 million people, or 4.7 per 100,000) (*40*) and the National Association of County & City Health Officials (NACCHO) recommended staffing levels during epidemics of 30 tracers per 100,000 people (*41*). We set the tracing delay to one day to be consistent with Washington state’s goal of notifying 80% of contacts within 48 hours (*42*), and use the King County Phase 2 Application to compute the expected rate of initial contact tracing interviews and follow-up notifications. Over a two-week period, 22 staff members contacted 336 individuals for initial interviews and 941 for close contact notifications, or approximately 1 initial interview and 3 notifications per day per staff member (*40*).

**Fig. 6.**
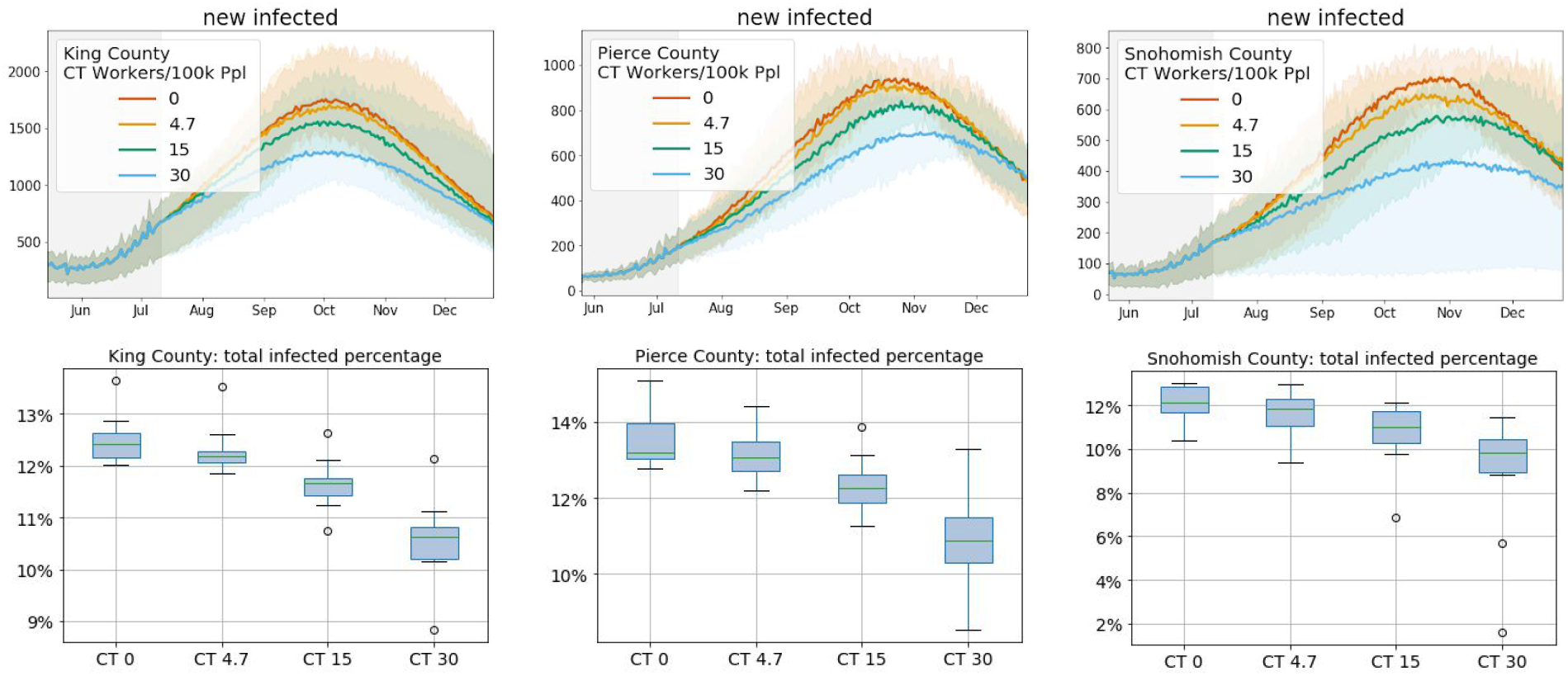
Estimated effect of manual contact tracing on new infections (top) and total infected percentage (bottom) at various staffing levels per 100k people in King, Pierce, and Snohomish counties between July 11, 2020 and December 25, 2020.

Manual tracing with the full desired staffing levels of 15 workers per 100,000 people is able to affect the epidemic trend in all three counties, but has a significantly smaller effect at current staffing levels (Fig. 6). Unsurprisingly, the impact for a given level of staffing is dependent upon the current epidemic trend, reinforcing the need for concurrent interventions to effectively manage the epidemic.

Additionally, we compare the performance of exposure notification to manual contact tracing to establish similarities between relative staffing level and exposure notification adoption and to verify an additive effect of concurrent manual tracing and exposure notification.

We see improvements in all cases when combining interventions (Fig. 7). In all three counties, exposure notification has a stronger effect at the given staffing and adoption levels, but adding either intervention to the other results in reduced infections, albeit to different extents based on the trend of the epidemic. This suggests that both methods are useful separately and combined, even if they do not explicitly coordinate.

**Fig. 7.**
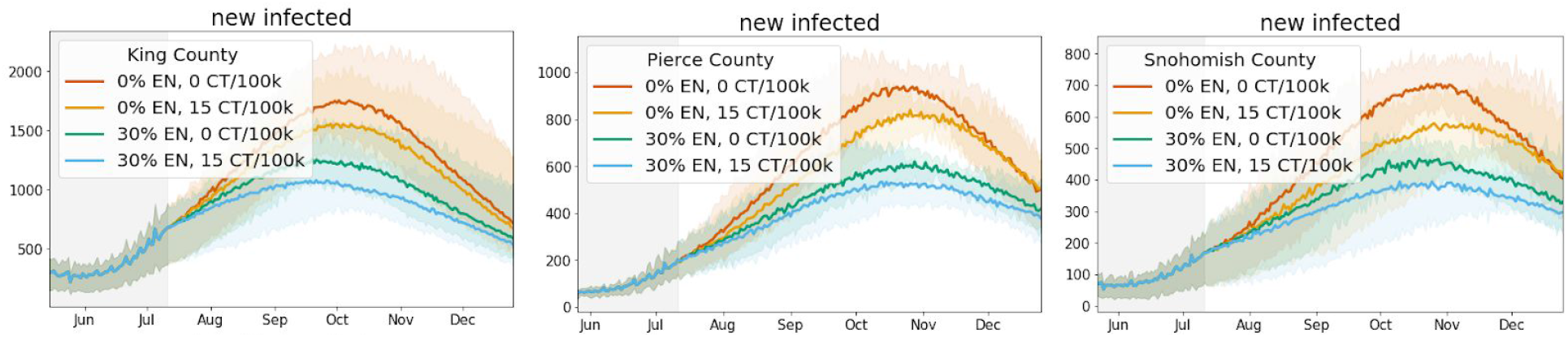
Comparison between manual contact tracing (CT) at the recommended staffing level and exposure notification (EN) at 30% adoption in King, Pierce, and Snohomish counties.

### Concurrent interventions under behavioral changes

While the results shown above suggest that the interventions are effective in suppressing the COVID-19 epidemic to various degrees, in practice, health organizations will implement multiple intervention strategies simultaneously to try to curb the spread of the virus while also allowing controlled reopenings. Therefore, we also study the combined effect of concurrent interventions including digital exposure notification, manual contact tracing, and social distancing (Fig. 8). We model social distancing as a function of infectiousness of interactions in the random and occupation networks, where increasing social distancing decreases the relative transmission likelihood on a network by a multiplicative factor relative to their values as of March 1, 2020 (i.e., before broad-based social distancing and mobility reductions). For example, social distancing of 1.7x is equivalent to multiplying the relative transmission by 1 / 1.7 = 0.6. Note that this does not change the number of person-to-person interactions, but rather the likelihood of transmission of any individual encounter, which may be affected by factors other than physical distancing such as mask usage, improved hygiene, use of personal protective equipment, etc.

**Fig. 8.**
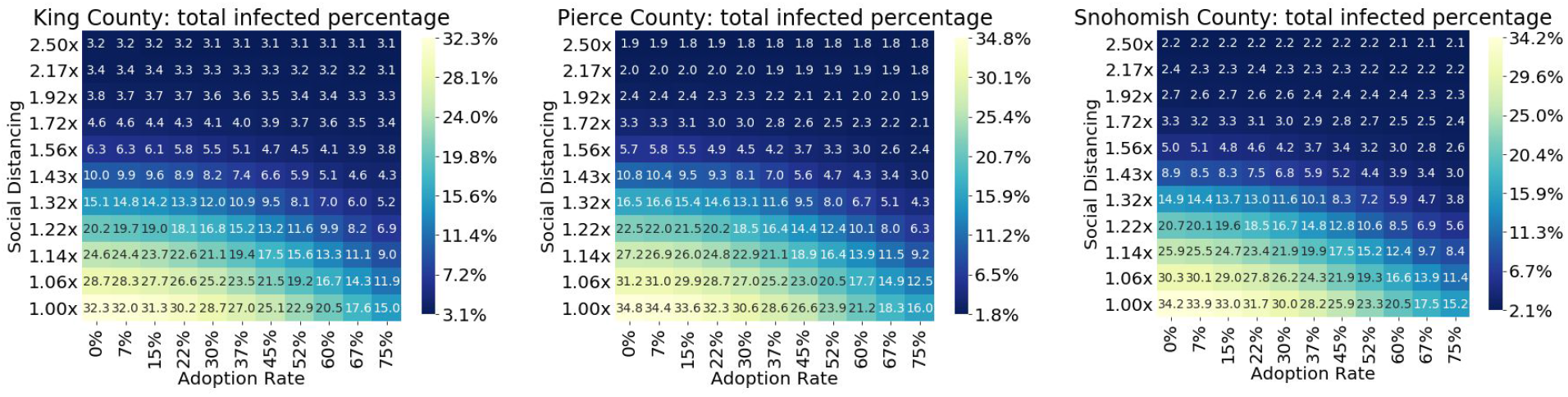
Estimated total infected percentages between July 11 to December 25, 2020 for King, Pierce, and Snohomish counties as a function of simultaneous social distancing and exposure notification app adoption. Social distancing is expressed as the infectiousness of random and occupation network interactions, relative to their initial values (i.e., before broad-based social distancing and mobility reductions).

Next we examine the effects of combined NPIs under various “reopening” scenarios by gradually increasing the number of interactions in every interaction network, including households, workplaces, schools, and random networks. Specifically, we increase these interactions by a given percentage from the levels as of July 11, 2020 (0% reopen) up to the initial levels at March 1, 2020, at the very start of the epidemic (100% reopen). Given the average number of interactions *i* for network *n* at the end of the baseline as *i_b,n_* and before the lockdown as *i*_0_*_,n_*, the network reopening percentage *p* (in 0-100%) defines the current relative interactions under reopening *i_c,n_* as

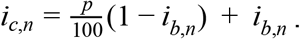

The increase in new infections from a 10-20% reopening are balanced by 22-37% exposure notification app adoption, although the effect varies by county (Fig. 9). This shows that limited additional reopenings may be possible after introducing exposure notification alongside existing fully staffed manual tracing (15 staff per 100,000 people), but that social distancing remains an important measure under these circumstances. Additionally, there is an increased effect to adding exposure notification under greater reopening scenarios. As an example, we plot some primary metrics for a 50% network reopening and see significant reductions in nearly all metrics at even 30% adoption (Fig. 10).

**Fig. 9.**
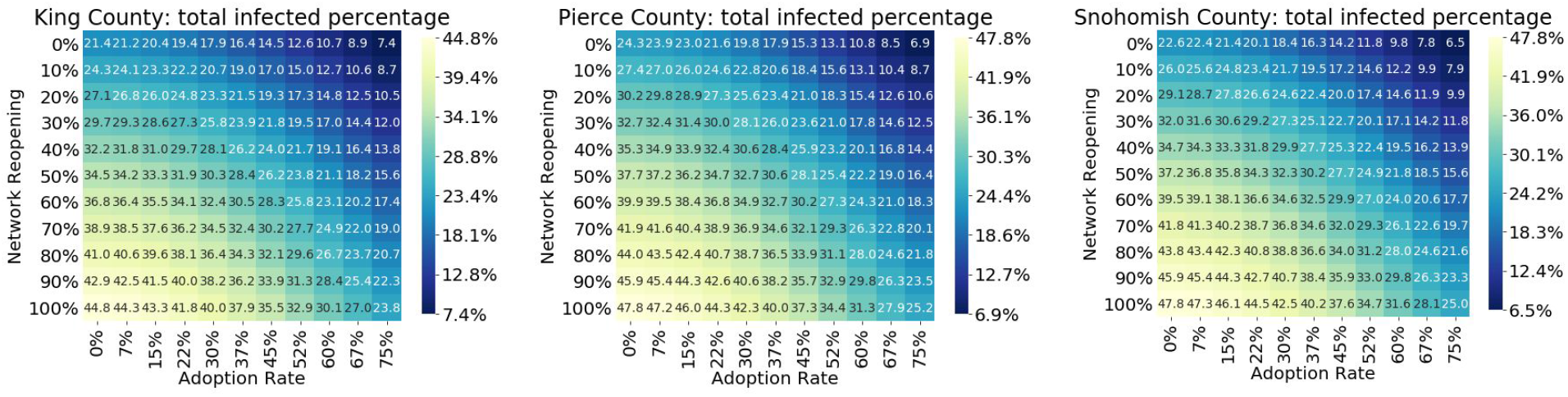
Estimated total infected percentage as a function of simultaneous network reopening and exposure notification app adoption rates, assuming fully staffed manual contact tracing (15 workers per 100,000 people).

**Fig. 10.**
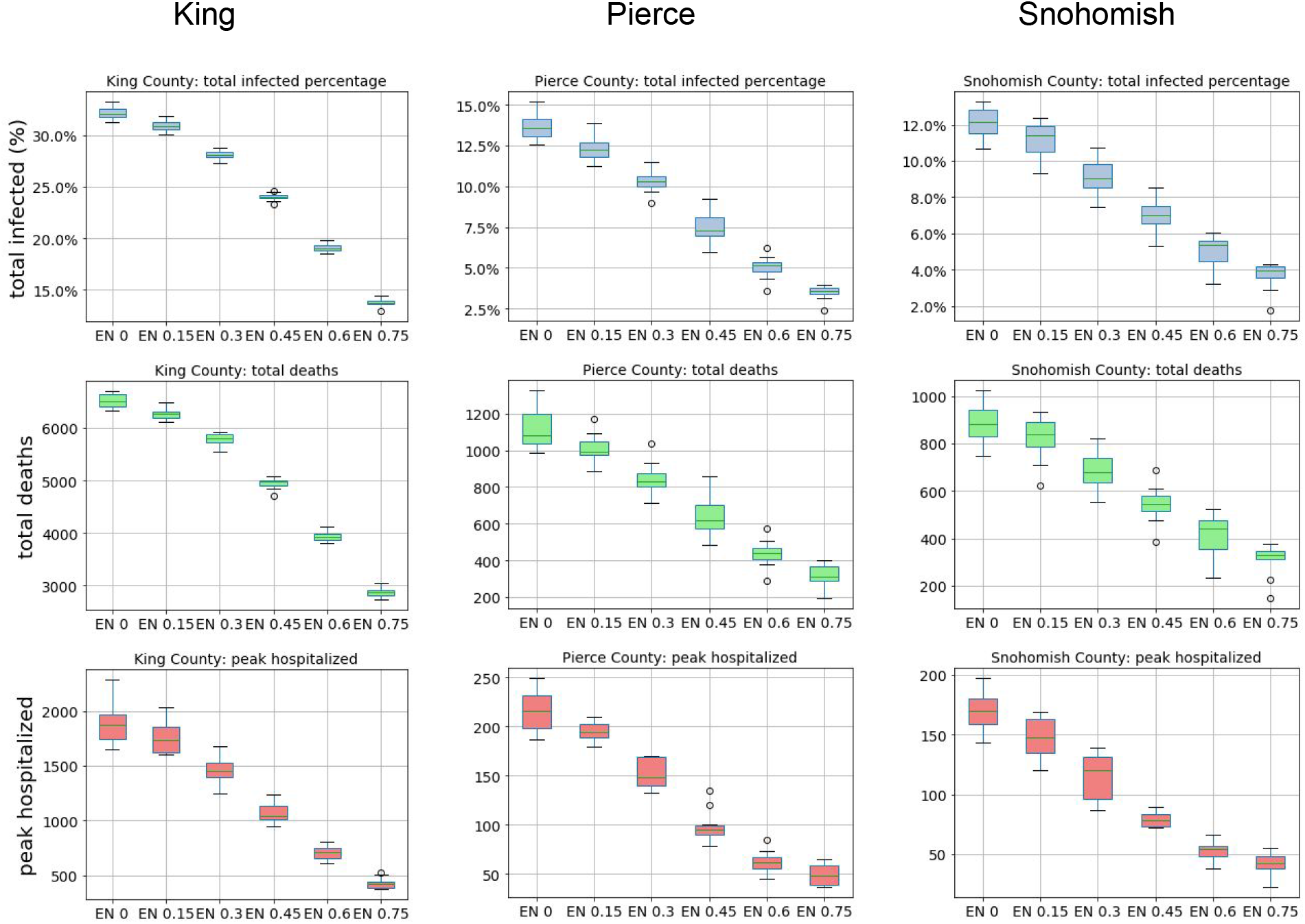
Estimated total infected percentage, total deaths, and peak hospitalized under a 50% reopening scenario (an increase of 50% of the difference between pre-lockdown and post-lockdown network interactions) at various exposure notification adoption rates for King, Pierce, and Snohomish Counties, assuming no change to social distancing β(t) after the baseline and 15 manual contact tracers per 100k people.

As part of the Washington State Department of Health’s “Safe Start” plan, a key target metric to reopen Washington is to reach fewer than 25 new cases per 100,000 inhabitants over the prior two weeks (*42*). Here, we examine how many days it would take to reach that target under the combined NPIs. With the recent spike in cases, the trajectory for reaching these targets without renewed lockdowns is out of the range of the simulations. Therefore, to show the relative benefits of the NPIs, we introduce an artificial renewed lockdown at the mobility levels averaged over the month before the Phase 2 reopenings (Phase 1.5 for King County) that occurred on June 5, 2020. Using this averaged mobility from May 6 to June 5, 2020, we model the relative effects of manual tracing and exposure notification on the Washington Safe Start key metric.

We find that for all three counties, manual contact tracing at the recommended staffing levels combined with an exposure notification app can significantly reduce the amount of time it takes to achieve this metric (Fig. 11). Under the recommended standard for manual tracing, adding exposure notification at 30% adoption results in reaching the target in 92%, 87%, and 85% of the time versus no exposure notification for King, Pierce, and Snohomish counties respectively. At the reduced levels of 4.7 tracers per 100,000 population, the target is reached in less than 83% and 88% of the time for King and Snohomish respectively, although the exact ratio can not be calculated as the metric is not achieved in the baseline simulation.

**Fig. 11.**
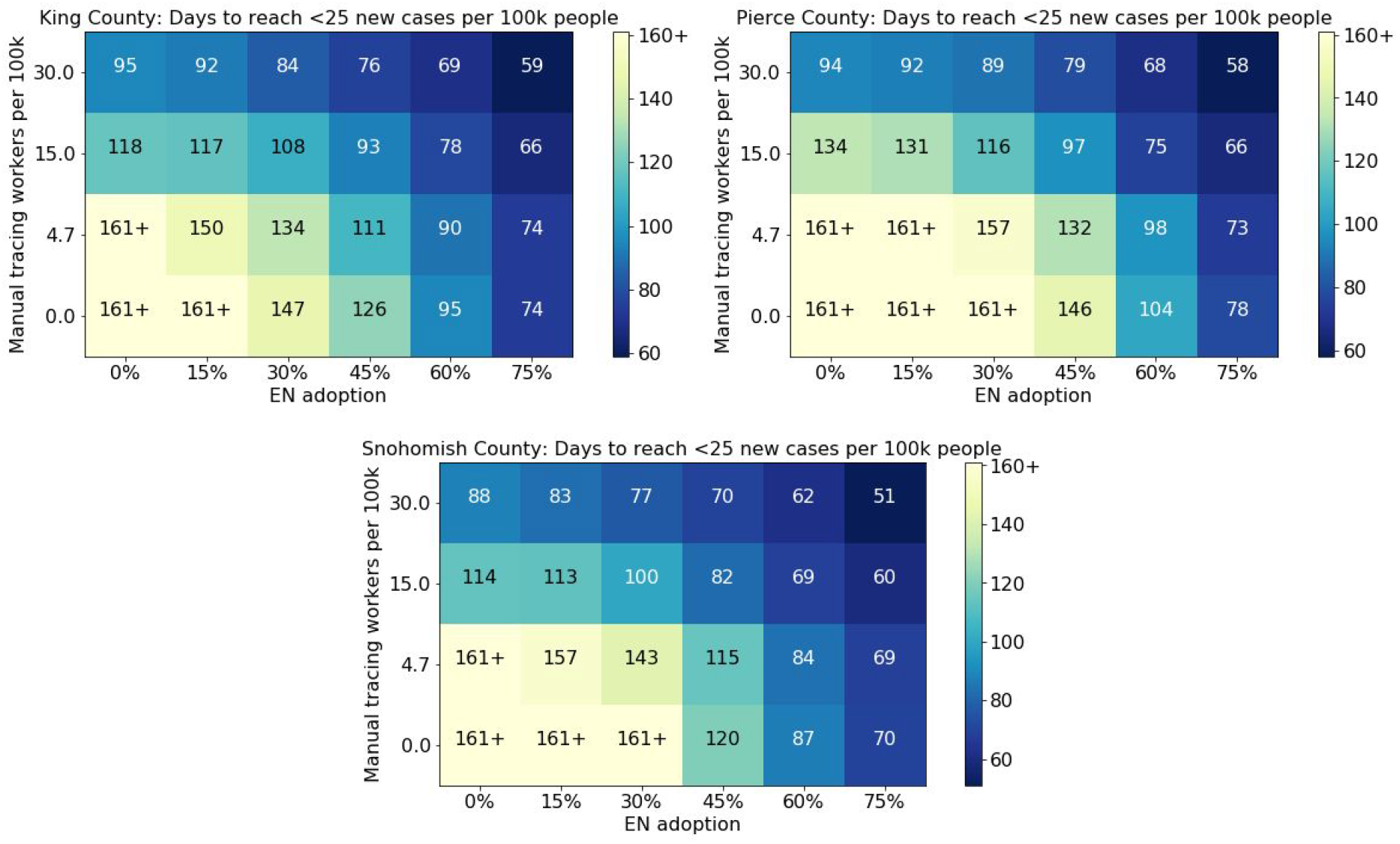
Estimated number of days from July 11, 2020 for King, Pierce, and Snohomish counties to reach the Washington state goal of fewer than 25 new cases per 100,000 people over the trailing 14 days, as a function of manual tracing workforce capacity and exposure notification app adoption, given a renewed lockdown to the average level over the month before June 5th.

## Limitations and Assumptions

Our individual-based modeling approach attempts to simulate the behavior of humans in a complex environment, in order to better understand the relative effects of different levels of intervention. While we have attempted to add realistic elements and calibrate it with the best available data, it still represents a dramatic simplification of the real world. Choices and simplifications made surrounding the behavior of the individuals, their movements in the world, disease dynamics, and many others, mean that the results should be viewed as an exploration of possible outcomes, not a prediction (*43*).

A more specific limitation in our work is that we modeled each county separately without cross-county interactions. In particular, we did not model how cross-county human movement contributes to disease spreading. We plan to explore this effect in our future work.

Our simulations assume that it takes 2 days from symptom onset to receive a COVID-19 test result and we acknowledge that this is a key assumption underlying our findings. Ferretti et al. (*44*) showed that the delay between the initial exposure to case confirmation, notification, and quarantine has a significant impact on the efficacy of the intervention. Rapid testing protocols can shorten the time between symptom development and case confirmation, and are essential for epidemic control (*20*).

We used published COVID-19 mortality data to calibrate model parameters. While the death count is arguably a good proxy to the true infection numbers, the published mortality data are scarce and noisy in small counties, resulting in the difficulty of modeling those counties with accuracy.

The synthetic occupation networks are based on the latest employment data corresponding to the fourth quarter in 2019 (*31*). Since the beginning of the pandemic, the size and structure of occupation networks may have changed compared to the latest available data.

In our work we used the mobility data along with a changepoint to model time-varying infection rates. While the changepoint vector models the net effect of various latent factors, it may be limited when multiple change points or more complex latent factors exist. The derived time-varying infection rate is homogeneously distributed to the random network and occupational networks. This is an approximation to the reality where the change may vary on different networks.

## Related Work

The compartmental modeling approach (*45*) (*46*) (*47*) has been widely used for epidemic study. This approach segments the total population by subgroups according to the disease progression stage and models the transmission of stages with differential equations. SEIR (susceptible-exposed-infected-recovered) (*48*) (*49*) (*50*) (*44*) is a common type of compartmental model used to study COVID-19 spread. However, this approach is not suitable for studying the impact of individual level interventions like exposure notification apps because they characterize the disease dynamics at a population-level.

In contrast to the compartmental model, the individual-based modeling approach (*13, 18, 19, 51–62*) simulates the infectious disease progression of individuals and can consider demographics, social interactions, and the environment. These individual-based models can predict the spread of COVID-19 in multiple countries by fitting the stochastic model of disease progression and human interactions from historical data. However, the impact of additional interventions such as digital exposure notification is unexplored.

In (*63*) (*64*), disease transmission is modeled by a stochastic process to fit the reproduction number of the total population. However, manipulating the reproduction number by real contact tracing actions can be challenging as it is subject to human interaction patterns, adoption rate, and many other types of interventions. This model lacks the characteristics of individuals as it uses the mean field theory to approximate the total population. (*65*) (*66*) (*67*) study contact tracing by situating individuals randomly in a space and mimicking human contacts by the individual’s collision from the spatial movement. While this spatial individual-based model reveals promising results in virus spread in relatively small and closed areas, such as public buildings (*68*), and cruise ships (*69*), the ad-hoc assumptions in individual mobility patterns are not suitable for studying the impact of contact tracing in the scale of a city. (*70*) introduces the spatial temporal model which has more realistic mobility patterns. However, the spatial movement used in these models is a simplification of contact tracing which lacks the individual interactions among family members, workmates and from random activities. The effectiveness of manual and digital contact tracing is discussed in (*71*) through empirical contact data collected from the work related network at a small scale, without considering virus spread among family members and other random interactions. The references (*57*) (*72*) are the closest to ours, but they do not cover the joint impact of manual and digital contact tracing. In addition, model calibration is missing in their case studies. In contrast, OpenABM-Covid19 (*22*) simulates concurrent manual contact tracing and digital exposure notification interventions over interaction networks at a large scale.

## Discussion

In this study we conducted a model-based estimation of the potential impact of a digital exposure notification app in Washington state. OpenABM-Covid19 simulates interactions among synthetic agents in various small-world networks, representing households, workplaces, schools, and random interactions. Interactions in those networks can result in COVID-19 transmission and are recalled to simulate different tracing interventions, including “manual” contact tracing or digital exposure notification, such as the recently released Apple and Google Exposure Notifications System (ENS). We calibrated our model using real-world data on human mobility and showed how it can accurately match epidemiological data in Washington state’s three largest counties, King, Pierce, and Snohomish.

Similar to Hinch et al.’s report on digital contact tracing in the UK (20), we found that a digital exposure notification app can meaningfully reduce infections, deaths, and hospitalizations in these Washington state counties at all levels of app uptake, even if only a small fraction of the eligible population participates. We also showed how digital exposure notification can be combined with manual contact tracing at the recommended levels to further suppress the epidemic, even if the two interventions do not explicitly coordinate. Our simulations showed that the simultaneous deployment of both interventions can help these Washington counties meet the key incidence metric defined by the Safe Start Washington plan before December, 2020. The potential overall effect of digital exposure notification seems to be greater than even optimal levels of manual contact tracing, likely because of its ability to scale and better identify random interactions.

We also found that quarantine rates, which contribute to the social and economic cost of these interventions, scale sublinearly with app adoption, meaning that in some cases there are fewer people quarantined even though a greater fraction of the population is participating in the app. We credit this effect to the success of the app at suppressing the epidemic at high levels of adoption. Given a longer simulation time horizon we may see a similar effect even at the lower levels of app adoption. Health authorities may consider this when appealing to the public by explaining how greater rates of collective participation may reduce the severity of the epidemic while also minimizing or reducing the need for quarantine.

Finally, we looked at the combined effects of digital exposure notification and manual tracing in the context of different reopening scenarios, where mobility and interaction levels increase to the pre-epidemic levels. Our results suggest that both interventions are helpful in counterbalancing the effect of reopening, but are not totally sufficient to offset new cases except at very high levels of adoption and manual tracing staffing. As a result we believe that continued social distancing and limiting person-to-person interactions is essential. Future work is needed to study targeted reopening strategies, such as reopening specific occupation sectors or schools, or more stringent social distancing interventions in places that do reopen.

Looking ahead to future work, we are considering the question of coordination between different regions when deploying digital exposure notification as part of a suite of non-pharmaceutical interventions. The United States has seen a highly spatially varied response to the COVID-19 pandemic, with significant consequences to epidemic control (*73*). Under the conditions of varying cross-county and cross-state flows, we seek to quantify the empirical efficiency gap between coordinated and uncoordinated deployments and policies around testing, tracing, and isolation in which a digital exposure notification system can aid. In particular, the beginning of such cross-state collaborations is evident in the consortia of state governments such as the Western States Pact and a multi-state council in the northeast, both working together to coordinate their responses. We expect that coordinated deployments of digital exposure notification applications and public policies may lead to more effective epidemic control as well as more efficient use of limited testing and isolation resources.

## Data Availability

The OpenABM-Covid19 code is publicly available at (22) and licensed under the GNU General Public License v3.0.

https://github.com/BDI-pathogens/OpenABM-Covid19

https://www.google.com/covid19/mobility/

https://www.nytimes.com/interactive/2020/us/coronavirus-us-cases.html

## Acknowledgements

We thank Luca Ferretti, Chris Wymant, Kevin Murphy, Carla Bromberg, Karen Smith, Kathryn Rough, Michael Howell, and Andrea Stewart for their helpful guidance.

## Funding

C.F. is funded by the Li Ka Shing Foundation.

## Author contributions

Conceptualization: S.O., P.E., A.W., Y.S., M.R., C.F. Methodology: R.H., C.F., W.P., A.N., L.A.-D., D.B., M.A., N.W., L.L. Data acquisition: M.D., M.A., S.O., N.W. Writing, original draft: M.A., S.O., N.W., P.E., Z.C., L.L. Writing, review, and editing: all authors.

**Data and materials availability:** the OpenABM-Covid19 code is publicly available at (22) and licensed under the GNU General Public License v3.0.

## Competing interests

M.A., N.W., L.L., A.W., P.E., Y.S., M.R., M.D., Z.C., S.O. are employees of Alphabet, Inc., a provider of the Exposure Notification System; no other relationships or activities that could appear to have influenced the submitted work.

## Footnotes

1 We exhaustively searched from 3.0-7.0 for the infectious rate parameter and 35 day period for the infection seed date.

